# Insomnia, sleep apnea, and incidence of hypertension and cardiovascular disease

**DOI:** 10.1101/2025.06.02.25328832

**Authors:** Allison E. Gaffey, Matthew M. Burg, Henry K. Yaggi, Kaicheng Wang, Cynthia A. Brandt, Sally G. Haskell, Lori A. Bastian, Tiffany E. Chang, Allison Levine, Melissa Skanderson, Andrey Zinchuk

## Abstract

**Introduction:** Comorbid insomnia and obstructive sleep apnea (OSA, i.e., COMISA) are associated with cardiovascular disease (CVD) among older adults. It is unknown how the comorbidity is related to cardiovascular risk among younger military Veterans, who show a greater risk for hypertension and CVD than non-Veterans, and if associations differ by sex. Thus, we examined whether COMISA is associated with incident hypertension and CVD risk in younger men and women Veterans.

**Methods:** The cohort included post-9/11 Veterans who enrolled in Veterans Health Administration care from 2001 to 2021. Administrative and electronic health record data were merged. Insomnia and OSA were defined by 2 outpatient International Classification of Diseases, 9 or 10 diagnoses. Hypertension was defined by ≥2 outpatient-coded diagnoses or ≥1 antihypertensive medication fill. CVD was defined by 1≥ inpatient or ≥2 outpatient diagnoses. Time-varying Cox proportional hazard models were adjusted for demographics, behavioral, and clinical factors and conducted overall and by sex.

**Results:** Analyses included 937,598 Veterans (12% women; median age: 41 years). Greater hypertension risk was observed overall (adjusted hazard ratio [aHR]:2.43: 95%CI:2.36-2.50), for men (aHR:2.09, 95%CI:2.02-2.16) and women with COMISA (aHR:2.20, 95%CI:2.00-2.42), insomnia only (aHRs:1.27-1.44), and OSA only (aHRs:2.00-2.26) versus no sleep disorder. For incident CVD, COMISA was again associated with risk overall (aHR:3.81, 95%CI:3.64-3.99), in men (aHR:3.81, 95%CI:3.63-4.00), and women (aHR:3.44, 95%CI:2.98-3.98), as were insomnia (aHRs:1.36-1.37) and OSA (aHRs:3.32-2.62).

**Conclusions:** For post-9/11 Veterans, COMISA was associated with the greatest risk of hypertension and CVD. Identifying disordered sleep among men and women should be a cardiovascular prevention priority.

## INTRODUCTION

Insomnia and obstructive sleep apnea (OSA) are the most common sleep disorders and often present concurrently, known as COMISA.^1^ Importantly, insomnia and OSA both contribute to the risk for hypertension, cardiovascular disease (CVD), and related mortality.^2–9^ As risk for CVD increases with age, identifying modifiable risk factors for hypertension and CVD may inform preventive strategies in younger adults. Still, most studies are conducted in middle-aged and older adults, and little is known about associations of insomnia, OSA, and COMISA with hypertension and cardiovascular risk among younger populations. Compared to non-Veterans, Veterans are particularly vulnerable to sleep disorders, hypertension, and CVD,^10–13^ and in whom CVD occurs at younger ages.^12^ Resolving these questions is valuable for the selection and timing of approaches to hypertension management and CVD prevention, with sleep disturbances as a modifiable target.

The cardiovascular risk associated with insomnia, OSA, and COMISA may also differ by sex. Men and women exhibit differences in the prevalence of common sleep disorders; specifically, OSA is more prevalent among men until women reach menopause when the sex difference narrows,^14^ while insomnia is more prevalent among women.^15,16^ Among military personnel, sex-specific characteristics of patients with COMISA have also been reported, with a greater apnea-hypopnea index among men than women and women endorsing greater symptoms of anxiety, posttraumatic stress disorder, and nightmare disorder than men.^17^ Hypertension risk is also affected by age and sex. The risk for hypertension is greater for younger and middle-aged men than women, although postmenopausal women and same-aged men show a similar prevalence of hypertension.^18^ Thus, sex may be an important factor in hypertension and cardiovascular risk from COMISA, but remains underexplored.

The primary objective of this investigation was to determine the overall and sex-specific associations of insomnia, OSA, and COMISA, with risk of incident hypertension and CVD in a nationwide sample of primarily younger- and middle-aged men and women Veterans. We hypothesized that for both men and women, insomnia, OSA, and COMISA would each be independently associated with a greater risk of hypertension and CVD than having no sleep disorder. Second, for both men and women that the effect of COMISA would be larger than those attributable to the independent effects of insomnia or OSA alone. Finally, due to the evidence that sleep disorders may be more often diagnosed in the VA for men, we expected that the effects of COMISA on hypertension and CVD risk would be greater for men than women.

## METHODS

Data used for these analyses are unavailable for distribution by the authors as they are covered by a Department of Veterans Affairs proprietary data use agreement.

### Data Sources

The VA’s Defense Manpower Data Center – Contingency Tracking System was used to identify eligible patients based on their dates of military discharge and establishment of VA care. This list was merged with national VA Corporate Data Warehouse electronic health record (EHR) data, which consists of all vital signs, procedural administrative, pharmacy, and diagnostic records (based on the International Classification of Diseases, Ninth Revision, Clinical Modification, and Tenth Revision [ICD-9-CM and ICD-10-CM] codes and dates), from Veterans’ VA healthcare visits. ICD-9-CM and ICD-10-CM criteria were used to determine all diagnoses (see eTable S1 for codes).

### Study Population

The Women Veterans Cohort Study (WVCS) is a nationwide, prospective examination of men and women Veterans who served during conflicts in Iraq and Afghanistan (i.e., post-9/11), were discharged from service October 1, 2001-September 30, 2021, and whose first Veterans Health Administration (VA) outpatient medical visit was before September 30, 2021. Patients were excluded if they were deceased during the period of follow-up, diagnosed with insomnia, OSA, hypertension or CVD before establishing VA care, if they had a history of a psychiatric disorder (i.e., bipolar or schizophrenia), missing demographics or covariates, or missing a date of follow-up care. Those with a prior history of CVD were also excluded for analyses with the hypertension outcome. The study characteristics are reported based on the Strengthening the Reporting of Observational Studies in Epidemiology (STROBE) guidelines (eTable S2).^19^

### Exposures: Insomnia, Sleep Apnea, COMISA

Patients were categorized as having insomnia or OSA if they had ICD 9/10 diagnostic codes on ≥2 outpatient visits. Using these diagnoses, four groups were created: insomnia only, OSA only, comorbid insomnia and OSA (COMISA), and neither diagnosis. The date of being coded for these diagnoses was also determined and used to calculate the time to each outcome event.

### Outcomes: Incident Hypertension and CVD

The primary outcome of incident hypertension was defined by an ICD 9/10 diagnosis of hypertension coded on at least 2 outpatient visits or a filled prescription for any antihypertensive medication (i.e., angiotensin-converting enzyme inhibitors, angiotensin II receptor blockers, beta-blockers, calcium channel blockers, or diuretics). The secondary outcome of incident CVD was defined as the presence of heart failure, peripheral vascular disease, atrial fibrillation, myocardial infarction, or ischemic stroke diagnoses on at least 1 inpatient discharge or 2 outpatient visits. This approach aligns with past investigations of CVD in VA EHR datasets.^20^ A third, exploratory outcome was having ≥2 outpatient blood pressure (BP) readings that exceeded the threshold of 140mmHg for systolic blood pressure or 90 mmHg for diastolic blood pressure per AHA/ACC guidelines (stage II hypertension).^21^

### Other Descriptive Variables and Covariates

Patients’ marital status, the availability of additional non-VA health insurance at baseline, baseline BP, and history of sleep assessment and treatment with medication for insomnia (i.e., doxepin, eszopiclone, ramelteon, suvorexant, temazepam, trazodone, zaleplon, zolpidem, or zolpidem tartrate) or positive airway pressure (PAP) therapy (e.g., E0601, E0470, E0561, A7027), were also used to characterize the sample. The *a priori* covariates were recorded at each patient’s baseline (i.e., when they established VA care and completed their first outpatient visit) or during the first documentation during VA follow-up. These variables included demographics: age, sex (women vs men [reference]) race/ethnicity (non-Hispanic Black, Hispanic, Other, non-Hispanic White [reference]), rurality (urban [reference], rural, very rural); behavioral factors: smoking history (never [reference], current, past), body mass index (BMI; categories of underweight/normal [reference], overweight, obese), and history of an alcohol or substance use disorder (ever diagnosed vs. never [reference]); and other clinical risk factors (ever diagnosed or screened positive vs. never [reference]): Type 2 diabetes, dyslipidemia, major depressive disorder (MDD), generalized anxiety disorder (GAD), and posttraumatic stress disorder (PTSD). For sensitivity analyses, a variable representing VA healthcare utilization was derived from the number of patients’ primary care visits during the first 2 years of VA care, as used in past analyses of VA EHR data.^20^

### Statistical analysis

Descriptive and bivariate statistics (chi-square for proportions, ANOVA for normally distributed variables, one-way ANOVA and Mann-Whitney for non-normally distributed variables) were used to present univariate data and to compare the relations of demographic, behavioral, and clinical factors by sex. The cumulative incidence of hypertension and CVD were then presented overall and for men and women separately, based on the number of cases per 1,000 person-years. Follow-up was censored at the data of hypertension or CVD diagnosis, or the date of a patients last VA encounter, based on the event that occurred first. Finally, time-varying Cox proportional hazard models were used to investigate the associations between sleep diagnoses, incident hypertension, and incident CVD, using patients without a past sleep diagnosis as the reference group.

Our primary analysis was modeling the associations of insomnia, OSA, and COMISA (vs. no sleep disorder) on hypertension incidence. Our secondary analysis included insomnia, OSA, and COMISA (vs. no sleep disorder) in relation to CVD incidence. Initial models included only the independent effects of insomnia, OSA, and COMISA in association with either hypertension or CVD. Subsequent models included adjustments for demographic, behavioral, and clinical risk factors. All models were conducted overall and then stratified by sex. To test the second hypothesis, if the COMISA effect was larger than each sleep disorder alone, additional analyses were conducted with OSA as the reference group to compare the additive effect of insomnia among those with COMISA. Finally, to determine if the effects of COMISA were greater for men than women, we conducted models with an interaction term by sex. Sensitivity analyses were also conducted to include healthcare utilization and to include outpatient BP readings in the hypertension definition. Missing values for covariates were assumed to be at random. Multiple comparisons were performed using the Bonferroni correction with *p* < 0.016 defined as statistically significant for each of the three pairwise comparisons. SAS version 9.4 (Cary, NC) was used for analyses.

## RESULTS

The final analytic sample included 937,598 patients (12% were women [n=113,381]), of which 13% had a diagnosis of insomnia, 20% had a diagnosis of OSA, and 14% were diagnosed with COMISA (Figure 1). The median age at baseline was 41 years; 61% of patients were non-Hispanic White, 50% were married, 72% lived in an urban location, and 35% had additional, non-government health insurance (Table 1). One-third were obese at baseline, and the most prevalent clinical disorders were PTSD, dyslipidemia, and MDD (42%-31%). Across the cohort, most patients completed one primary care visit in the initial two years of VA care, and the median follow-up period was 7 years. Of those with COMISA, 72% were first diagnosed with insomnia and 28% were first diagnosed with OSA.

**Figure 1.**
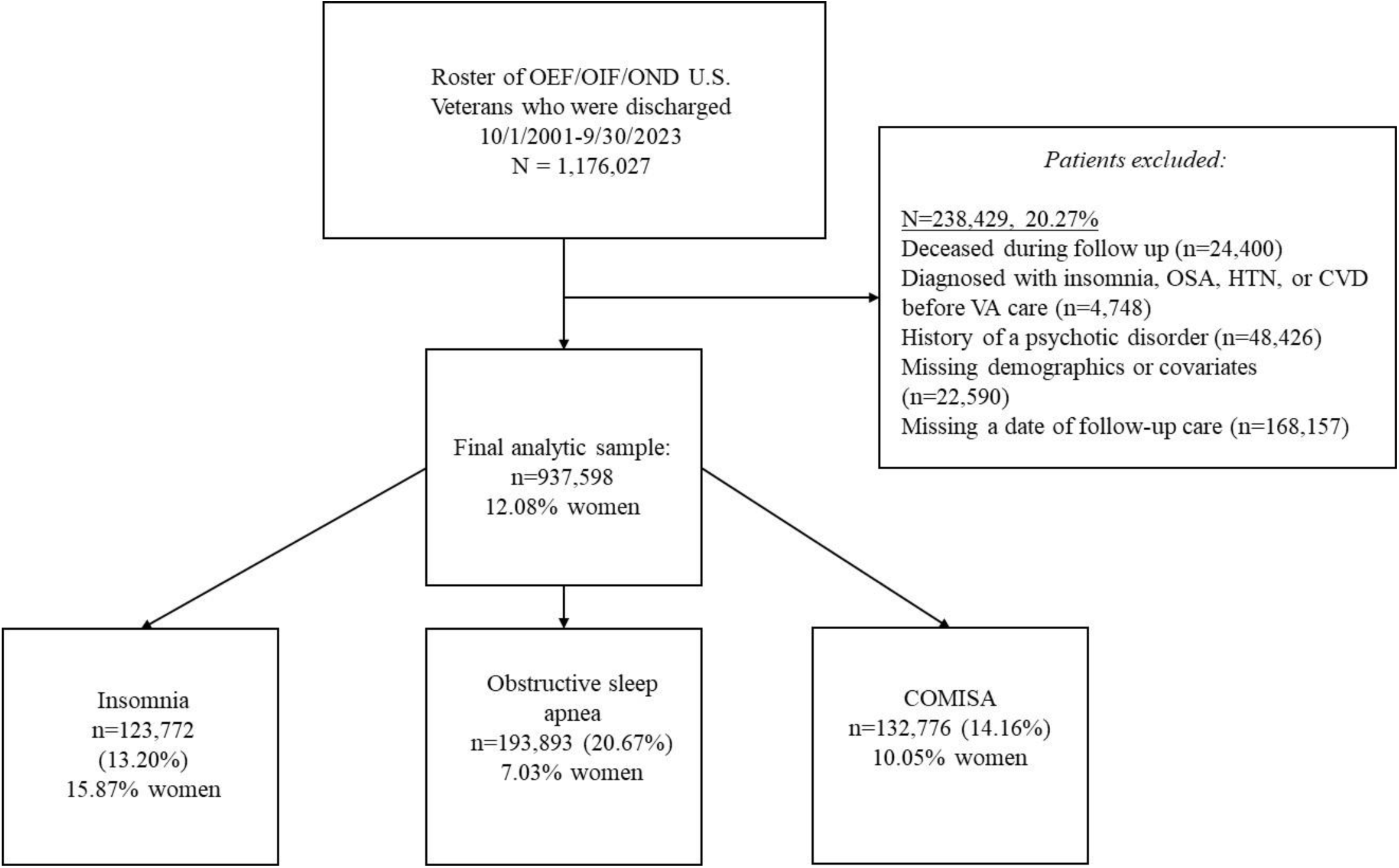
Flowchart of the study cohort.

**Table 1.** Demographic and health characteristics of the Veteran cohort.

|  | **Total**<br>**(n=937,598)** | **Men**<br>**(n=824,217)** | **Women**<br>**(n=113,381)** | **P** |
| --- | --- | --- | --- | --- |
| **Age, y (median)** | 41.0 (37.0,50.0) | 41.0 (37.0,50.0) | 41.0 (37.0,49.0) | <0.001 |
| **Age (years)** |  |  |  | <0.001 |
| **18-39** | 397985 (42.4) | 349811 (42.4) | 48174 (42.5) |  |
| **40-59** | 471757 (50.3) | 413846 (50.2) | 57911 (51.1) |  |
| **≥60** | 67856 (7.2) | 60560 (7.4) | 7296 (6.4) |  |
| **Race and ethnicity** |  |  |  | <0.001 |
| **Non-Hispanic Black** | 149805 (15.9) | 117710 (14.3) | 32095 (28.3) |  |
| **Hispanic** | 111222 (11.8) | 97359 (11.8) | 13763 (12.1) |  |
| **Non-Hispanic Other** | 96164 (10.2) | 83440 (10.1) | 12724 (11.2) |  |
| **Non-Hispanic White** | 580407 (61.9) | 525608 (63.8) | 54799 (48.3) |  |
| **Marital status** |  |  |  | <0.001 |
| **Married** | 475418 (50.7) | 431537 (52.4) | 43881 (38.7) |  |
| **Divorced** | 147038 (15.6) | 120724 (14.7) | 26314 (23.2) |  |
| **Never married** | 267811 (28.5) | 231192 (28.1) | 36619 (32.3) |  |
| **Rurality of residence** |  |  |  | <0.001 |
| **Urban** | 666907 (71.1) | 580231 (70.4) | 86586 (76.4) |  |
| **Rural** | 205924 (21.9) | 185820 (22.6) | 20104 (17.7) |  |
| **Highly rural** | 22316 (2.3) | 20319 (2.5) | 1997 (1.8) |  |
| **Additional health insurance** | 333466 (35.6) | 294922 (35.8) | 38544 (34.0) | <0.001 |
| **Baseline systolic BP, mmHg (median)** | 125 (117,134) | 126 (118,135) | 117 (109,126) | <0.001 |
| **Baseline diastolic BP, mmHg (median)** | 78 (70,84) | 78 (71,84) | 73 (67,80) |  |
| **Smoking status** |  |  |  | <0.001 |
| **Never smoker** | 388266 (41.4) | 320958 (38.9) | 67308 (59.4) |  |
| **Current** | 360297 (38.4) | 333641 (40.5) | 26656 (23.5) |  |
| **Former** | 25976 (17.3) | 146330 (17.8) | 16729 (14.8) |  |
| **BMI (median)** | 29.6 (26.3,33.3) | 39.7 (26.6,33.4) | 28.4 (24.7,32.7) | <0.001 |
| **BMI categories** |  |  |  | <0.001 |
| **Normal/underweight** | 115414 (12.3) | 91534 (11.1) | 23880 (21.1) |  |
| **Overweight** | 262955 (28.0) | 232660 (28.2) | 30295 (26.7) |  |
| **Obese** | 333323 (35.5) | 297593 (36.1) | 35730 (31.5) |  |
| **Alcohol abuse disorder** | 176325 (18.8) | 163963 (19.9) | 12362 (10.9) | <0.001 |
| **Drug use disorder** | 107519 (11.4) | 99560 (12.1) | 7959 (7.0) | <0.001 |
| **Diabetes** | 79638 (8.5) | 72030 (8.7) | 7608 (6.7) | <0.001 |
| **Dyslipidemia** | 380458 (40.5) | 347851 (42.2) | 32607 (28.8) | <0.001 |
| **PTSD** | 455926 (48.6) | 404732 (49.1) | 51194 (45.2) | <0.001 |
| **MDD** | 340981 (36.3) | 287086 (34.8) | 53895 (47.5) | <0.001 |

COMISA, HYPERTENSION, AND CARDIOVASCULAR DISEASE

|  |  |  |  |  |
| --- | --- | --- | --- | --- |
| **GAD** | 106694 (11.4) | 87003 (10.6) | 19691 (17.4) | <0.001 |
| **Insomnia** | 123772 (13.2) | 103380 (12.5) | 19695 (17.4) | <0.001 |
| **OSA** | 193893 (20.7) | 181443 (22.0) | 13721 (12.1) | <0.001 |
| **COMISA** | 132776 (14.2) | 120420 (14.6) | 13456 (11.9) | <0.001 |
| **First diagnosed with insomnia** | 72.1 | 73.0 | 63.6 | <0.001 |
| **First diagnosed with OSA** | 27.9 | 27.0 | 36.4 | <0.001 |
| **Healthcare utilization** | **Table** |  | **Women**<br>**(n=113,381)** |  |
| **Number of primary care visits within 2 years of cohort entry, median (IQR)** | 1 (0,4) | 1 (0,4) | 1 (0,5) | <0.001 |
| **Follow-up period, median (IQR)** | 7.2 (4.0,10.8) | 7.2 (4.0,10.7) | 7.5 (4.5,11.0) | <0.001 |
| **History of a sleep study** | 97057 (10.4) | 87162 (10.6) | 9895 (8.7) | <0.001 |
| **Prescription of insomnia medication** | 451471 (48.2) | 392487 (47.6) | 58984 (52.0) | <0.001 |
| **Prescription of CPAP** | 162376 (17.3) | 152102 (18.5) | 10274 (9.1) | <0.001 |
| **Incidence of hypertension** | 31.6 | 32.2 | 27.5 | <0.001 |
| **Incidence of CVD** | 14.3 | 14.3 | 14.9 | <0.001 |

Abbreviations: BP, blood pressure; BMI, body mass index; CPAP, continuous positive airway pressure device, GAD, generalized anxiety disorder; IQR, interquartile range; MDD, major depressive disorder; PTSD, posttraumatic stress disorder
Data are presented as N (%). All other data are presented as median (Q1, Q3). Group differences were examined using the chi-square test for categorical variables and the Kruskal-Wallis test for continuous non-normally distributed variables. Percentages may not add up to 100% due to rounding and missing data for a certain variable.

When comparing patient characteristics by sex, a greater percentage of men than women were non-Hispanic White (62% vs. 48%) and were married (51% vs. 27%). More men were diagnosed with a lipid disorder than women (39% vs. 27%), but fewer men were diagnosed with MDD (30% vs. 42%). A greater percentage of men had a diagnosis of OSA than women (32% vs. 21%), although there was a similar proportion of insomnia diagnoses by sex (23% vs. 24%). Among the group with COMISA, about 73% of men were diagnosed with insomnia before OSA compared to 64% of women. Among the COMISA group, while more men had a history of a sleep study or were prescribed CPAP than women (10% vs. 8% and 17% vs. 9%), slightly fewer men were ever prescribed insomnia medication (44% vs 49%).

### Risk for Incident Hypertension

Across the 20 years of VA follow-up, 310,048 patients developed hypertension (33%) and the median follow-up time was 7 years (IQR: 4,11). The incidence of hypertension was 31.6 per 1,000 person-years overall, 32.2 for men, and 27.5 for women. In unadjusted multivariable models, COMISA, insomnia alone, and OSA alone, were each associated with significantly greater risk for hypertension (Table 2; COMISA hazard ratio [HR]: 2.98, 95% CI:2.90-3.07, insomnia HR: 1.51, 95% CI:1.49-1.53), and OSA HR: 2.41, 95% CI:2.36-2.45) compared to patients without any sleep diagnosis. After adjustment for demographics, behavioral, and clinical factors, across the cohort, there was a greater risk for hypertension in those with COMISA (hazard ratio [aHR]: 2.43, 95% CI:2.36-2.50), insomnia (aHR: 1.44, 95% CI:1.42-1.46), and OSA (aHR: 2.26, 95% CI:2.22-2.30), compared to those without either diagnosis.

**Table 2.**
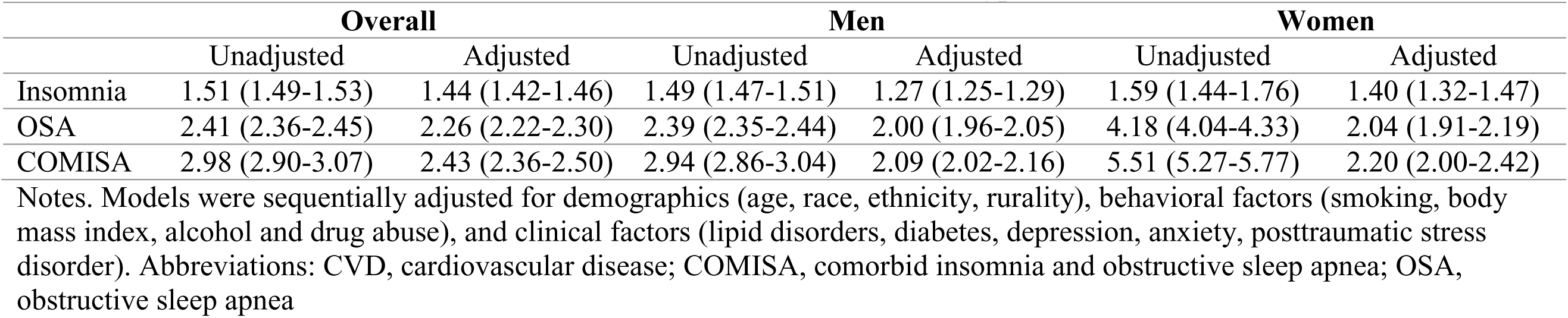
Associations of COMISA, insomnia, and OSA on risk for incident hypertension.

|  | Overall |  | Men |  | Women |  |
| --- | --- | --- | --- | --- | --- | --- |
|  | Unadjusted | Adjusted | Unadjusted | Adjusted | Unadjusted | Adjusted |
| Insomnia | 1.51 (1.49-1.53) | 1.44 (1.42-1.46) | 1.49 (1.47-1.51) | 1.27 (1.25-1.29) | 1.59 (1.44-1.76) | 1.40 (1.32-1.47) |
| OSA | 2.41 (2.36-2.45) | 2.26 (2.22-2.30) | 2.39 (2.35-2.44) | 2.00 (1.96-2.05) | 4.18 (4.04-4.33) | 2.04 (1.91-2.19) |
| COMISA | 2.98 (2.90-3.07) | 2.43 (2.36-2.50) | 2.94 (2.86-3.04) | 2.09 (2.02-2.16) | 5.51 (5.27-5.77) | 2.20 (2.00-2.42) |

Notes. Models were sequentially adjusted for demographics (age, race, ethnicity, rurality), behavioral factors (smoking, body mass index, alcohol and drug abuse), and clinical factors (lipid disorders, diabetes, depression, anxiety, posttraumatic stress disorder). Abbreviations: CVD, cardiovascular disease; COMISA, comorbid insomnia and obstructive sleep apnea; OSA, obstructive sleep apnea

Among men, there were significant unadjusted effects of COMISA (HR: 1.49, 95% CI:1.47-1.51), insomnia (HR: 2.39, 95% CI: 2.35-2.44), and OSA (HR: 2.94, 95% CI: 2.86-3.04) on risk of incident hypertension. For women, COMISA (HR: 1.59, 95% CI: 1.44-1.76), insomnia (HR: 4.18, 95% CI: 4.04-4.33), and OSA (HR: 5.51, 95% CI: 5.27-5.77) were each associated with greater hypertension risk. After adjusting the models for men, COMISA remained associated with hypertension risk (aHR:2.09, 95% CI:2.02-2.16), as did insomnia (aHR: 1.27, 95% CI: 1.25-1.29), and OSA (aHR: 2.00, 95% CI:1.96 -2.05). Among women only, the fully adjusted models were again significant for COMISA (aHR: 2.20, 95% CI:2.00-2.42), insomnia (aHR:1.40, 95% CI:1.32-1.47), and OSA (aHR: 2.04, 95% CI:1.91 -2.19).

Next, the reference group was changed from no sleep disorder to OSA to understand how much the relative risk of COMISA exceeded that of OSA. In the unadjusted analyses of the entire cohort and by sex, those with COMISA had over a 28-34% greater risk of hypertension than patients with OSA only. In fully adjusted models, overall, for men, and for women those with COMISA had a 17-26% greater risk of hypertension. Next, to examine the role of sex we included an interaction term – Sex x Insomnia x OSA – in the fully adjusted model of the whole cohort. While the overall interaction was not significant (*p*=0.817), interaction of Sex x Insomnia was associated with risk for hypertension (*p*s<0.001), indicating a greater effect of insomnia for women. Finally, sensitivity analyses were conducted to include adjustment for healthcare utilization, with results showing consistency with the primary analyses (eTables 3-5). Analyses that included BP readings of 140/90 mmHg in the definition of hypertension also aligned with the primary findings (eTables 6-8).

### Risk of Incident CVD

Overall, 3.2% of patients developed CVD. The cumulative incidence of CVD per 1,000 person-years was 14.3 overall, 14.3 for men, and 14.9 for women. In unadjusted analyses across the cohort, COMISA (HR: 5.54, 95% CI:5.28-5.80), insomnia (HR: 1.58, 95% CI:1.53-1.64), and OSA (HR: 4.21, 95% CI:4.07-4.36) were all associated with greater risk of all-cause CVD (Table 3). After adjusting for demographics, health behaviors, and clinical factors in the entire cohort, COMISA (aHR: 3.81, 95% CI:3.64-3.99), insomnia (aHR: 1.37, 95% CI: 1.32-1.41), and OSA (aHR: 3.32, 95% CI:3.21-3.43) were each associated with greater risk of all-cause CVD than that observed for patients without a disorder.

**Table 3.** Associations of COMISA, insomnia, and OSA on risk for incident all-cause CVD.

|  | Overall |  | Men |  | Women |  |
| --- | --- | --- | --- | --- | --- | --- |
|  | Unadjusted | Adjusted | Unadjusted | Adjusted | Unadjusted | Adjusted |
| Insomnia | 1.58 (1.53-1.64) | 1.37 (1.32-1.41) | 1.59 (1.44-1.76) | 1.36 (1.31-1.41) | 1.59 (1.54-1.65) | 1.36 (1.22-1.51) |
| OSA | 4.21 (4.07-4.36) | 3.32 (3.21-3.43) | 4.18 (4.04-4.33) | 3.36 (3.25-3.48) | 3.07 (2.72-3.46) | 2.62 (2.32-2.97) |
| COMISA | 5.54 (5.28-5.80) | 3.81 (3.64-3.99) | 5.51 (5.27-5.77) | 3.81 (3.63-4.00) | 4.45 (4.87-5.13) | 3.44 (2.98-3.98) |

Notes. Models were sequentially adjusted for demographics (age, race, ethnicity, rurality), behavioral factors (smoking, body mass index, alcohol and drug abuse), and clinical factors (hypertension, lipid disorders, diabetes, depression, anxiety, posttraumatic stress disorder); Abbreviations: CVD, cardiovascular disease; COMISA, comorbid insomnia and obstructive sleep apnea; OSA, obstructive sleep apnea

Men with COMISA (HR: 5.51, 95% CI:5.27-5.77), insomnia (HR: 1.59, 95% CI:1.44-1.76), and OSA (HR: 4.18; 95% CI:2.72-3.46) each had a greater, unadjusted risk of CVD, compared to men without a sleep disorder. These effects were similar among women, where the unadjusted effect was significant for COMISA (COMISA HR: 4.45, 95% CI:4.04-4.33; insomnia HR: 1.59, 95% CI:1.54-1.65; OSA HR: 3.07, 95% CI:2.72-3.46). For men, after adjusting for all covariates, COMISA was again associated with a greater risk of CVD (aHR: 3.82, 95% CI: 3.65-4.01), along with insomnia (aHR: 1.38, 95% CI: 1.33-1.43), and OSA (aHR: 3.38, 95% CI: 3.27-3.50). When modeling the adjusted effects of sleep disorders among women, the risk for CVD was greatest with COMISA (aHR: 3.40, 95% CI: 2.95-3.93), and was also significant for insomnia (aHR: 1.35, 95% CI:1.22-1.50), and OSA (aHR: 2.75, 95% CI: 2.44-3.10).

In unadjusted analyses with OSA as the comparison group, in the entire cohort, and for men and women separately, patients with COMISA had a 57%-90% greater risk of all-cause CVD than those without. After adjustment, patients with COMISA still had a 30%-53% greater CVD risk than those with OSA only. Next, the primary models were rerun with an interaction between sex and the sleep disorders. While there was a significant interaction of Sex x OSA in relation to risk for all-cause CVD (p<0.001), demonstrating the greater OSA-associated risk of CVD among men, the Sex x Insomnia x OSA interaction was not significant (ps=0.281). Finally, in sensitivity analyses adding healthcare utilization to the fully adjusted models, the effects remained overall, for men, and for women (eTables 9-11).

## DISCUSSION

Across this distinctive nationwide cohort of 1 million men and women who were served by the VA across 20 years, about 50% of Veterans were diagnosed with insomnia, OSA, or both disorders. Each of the sleep disorders was associated with increased risk of hypertension, beyond traditional factors, of which COMISA had the highest risk – over 2-fold. In addition to hypertension, this sleep-related risk was also associated with CVD, and the magnitude of risk for CVD was even greater than for hypertension – over 3-fold. Both men and women with COMISA seem to be at equivalent, adjusted risk for hypertension and CVD with a few exceptions (i.e., insomnia in relation to hypertension among women, OSA and CVD in men). However, these associations were robust for each group, included a long period of follow-up, and adjustment for multiple potential confounders and healthcare utilization, indicating that sleep disturbances and disorders are impactful risk factors that warrant the attention of cardiovascular medicine.

About 13% of the cohort were diagnosed with insomnia, 21% were diagnosed with OSA, and a separate 14% were diagnosed with both sleep disorders. In another sample of active duty military personnel referred for sleep assessment, 33% had only an insomnia diagnosis, 30% were diagnosed with OSA only, and a separate 37% were diagnosed with both.^22^ All the prevalence rates observed in the present study were below those observed in the sample of military personnel, indicating that active military have more issues with sleep than Veterans,^10^ or that sleep disorders may be underdiagnosed in the VA.^23,24^ Although Veterans commonly experience conditions that are comorbid with insomnia and OSA,^17,22^ and data from the general population show greater OSA rates in men and insomnia in women,^15,25^ insomnia was diagnosed equally across sexes, and OSA was more prevalent among men. Therefore, VA patients with insomnia may be overlooked or misdiagnosed with other comorbid conditions (e.g., depression), and OSA may be underdiagnosed in women Veterans, as observed more generally.^5,26^

We also found increasing hypertension risk among those with insomnia, OSA, and COMISA. Of the three sleep disorder groups, the risk associated with COMISA was greatest and appeared to reflect additive risk of the parent disorders. Insomnia, OSA, and COMISA were also each associated with increased risk of all-cause CVD compared to patients with no sleep disorder diagnosis, and COMISA-associated risk again showed the strongest effect. Because of the potential for inadequate diagnosis or misclassification of insomnia or OSA,^27^ the associations with hypertension and CVD could be more or less than observed. Analyses by sex also yielded interesting relations of sleep and cardiovascular risk. While the risks of hypertension and CVD often differed for men and women, as there were no significant interactions of both insomnia and OSA by sex, it seems the differences were not clinically meaningful. Of note, the associations were distinct from the influence of other demographic, behavioral, and clinical variables associated with hypertension and CVD risk, including age,^28^ race,^29^ weight,^30^ and psychiatric sequalae.^31,32^

The observed effects of COMISA align with previous evidence that co-occurring insomnia and OSA are associated with risks of hypertension, diabetes, and CVD.^7–9,33^ Notably, the two largest prior studies initially reported strong associations between COMISA and cardiovascular events, which were then attenuated after adjustment. In the Sleep Heart Health Study, 5803 adults (56% women) were followed for 11 years.^34^ While unadjusted results showed a twofold greater risk of CVD events in those with COMISA, the association was reduced and no longer significant after adjustment. Similarly, a retrospective review of electronic health records from 3951 sleep medicine patients (54% women) showed that COMISA was associated with a 3.6-fold greater risk of major adverse cardiovascular events, particularly in those aged 20-54.^33^ Again, this association became nonsignificant after accounting for clinical comorbidities. In our study, the effects ranged from 2.43 to 3.81 after adjustment. Two factors may explain the stronger associations observed in the present study. First, our clinical sample of Veterans likely had greater comorbidity burdens than the population-based samples in previous research, and the associated effects of disordered sleep on cardiovascular risk could also be more robust compared to those without the sleep disorders. Importantly, the effect sizes for COMISA in our study (e.g., for CVD, 3.44–3.81) exceeded those reported previously (1.71–1.88^7,8,35–39^), suggesting a potentially greater impact of disordered sleep on cardiovascular risk in Veterans. Second, differing study results may be partly due to sample ages: our cohort had a median age of 41 years (IQR:37,50), which is younger than those in other studies (e.g., mean age of 51,^38^ median of 55^8^). Past work indicated that the relation between OSA and incident hypertension may be stronger in younger to middle-aged adults,^40,41^ and it is plausible that COMISA similarly poses a greater cardiovascular risk in younger populations.^26,33^

Patients’ biological sex could influence the relationship between sleep disorders and cardiovascular risk, but there has been remarkably little inquiry into potential sex differences in these associations. While insomnia is more prevalent in women and OSA is more prevalent for men in the general population,^15,25^ this is the first known study to report COMISA prevalence separately for men and women Veterans, showing similar rates (12% and 10%). In contrast, another investigation of 1,096 non-Veterans who were referred to polysomnography showed that COMISA was more common in women than men (51% vs. 35%).^42^ However, age may modify these sex differences; among 860 patients (52% women), COMISA was more common in men ≤55 years than in same-aged women.^43^ For hypertension, the hazard associated with insomnia was greater for women than men, but hazards were not different for OSA or COMISA. When examining risk for CVD, the hazard of OSA was greater for men than women, but there was no difference for insomnia or COMISA. Yet, the magnitude of the observed differences was very small, suggesting there were no marked sex differences and that hypertension and CVD risk due to sleep disorders warrant clinical attention in both sexes, with two possible exceptions (i.e., insomnia and women’s risk for hypertension, and OSA in relation to men’s risk for CVD). Despite the empirical rationale for conducting sex-specific analyses, there have been few efforts to do so. In a 43-month prospective cohort study of 868 elderly patients (37% women), men with COMISA had a 2.8-fold increased risk of major adverse cardiovascular events, but no association was found for women.^44^ Another 12-year prospective study of 471 patients with type 2 diabetes (25% women) showed a 4-fold greater risk of CVD in those with COMISA, with no interaction with sex.^8^ To extend this sparse, inconsistent literature, investigating sex differences in COMISA presentation should be prioritized in future studies of CVD risk.

### Mechanisms of COMISA and CVD

There is likely a bidirectional association between the risk for OSA and insomnia.^1,45–47^ On one hand, ample evidence shows that OSA can predispose patients to insomnia if respiratory events and post-apneic waking periods are interpreted as difficulty staying asleep,^45,46^ which can increase patients’ alertness and anxiety about their sleep quality.^47^ Some patients treated with continuous positive airway pressure ventilation (CPAP) show greater sleep fragmentation,^47^ which may be linked to increased hypothalamic-pituitary-adrenal axis (HPA) activity and concurrent adverse metabolic changes.^47^ On the other hand, less evidence supports the theory that insomnia-related sleep deprivation could worsen OSA – e.g., increasing the AHI and reducing oxygen. Sweetman and colleagues have argued that insomnia could predispose individuals to a lower arousal threshold, thereby increasing patients’ vulnerability to OSA.^1^ Either causal association is plausible, and it is unknown if first developing insomnia vs. OSA then contributes differently to cardiovascular risk.

The pathophysiological consequences of COMISA on CVD may occur via several shared mechanistic pathways – sleep duration, fragmented sleep, and arousals – which could be stronger for those who are diagnosed with both sleep disorders.^48^ Independently and additively these factors promote autonomic dysfunction and systemic inflammation, followed by increased heart rate and blood pressure, myocyte toxicity, atherogenesis, and endothelial dysfunction. In addition to potential shared pathways, insomnia and OSA also have distinct effects on cardiovascular risk.

For insomnia, dysregulation of the HPA axis can alter insulin resistance and lipid levels, while OSA is associated with downstream changes in organ dysfunction, increased ventricular afterload, and aortic dilation.^48^ Scant data is available about sex differences in these pathways, but differences are possible. For example, it has been theorized that women with insomnia or OSA have a greater predisposition to adverse cardiovascular effects via inflammation. In one study of 323 women from the American Heart Association Go Red for Women Strategically Focused Research Network, self-reported symptoms of insomnia were associated with endothelial inflammation among women, although men were not included in the sample.^49^ Similarly in another population-based cohort of 479 adults, women with OSA showed greater systemic inflammation and high-sensitivity troponin-T than men.^50^ Yet, there is inadequate information about which COMISA-CVD pathways could be more influential for men compared to women.

### Risk Assessment and Management of COMISA

The need for routine sleep screening in patients with cardiovascular risk factors, and cardiovascular assessment in those with sleep disorders is becoming clearer. Our finding that the elevated CVD risk associated with COMISA is not solely attributable to OSA, underscores the importance of addressing both insomnia and OSA in clinical care. While treatment efforts for COMISA are expanding, they largely draw from approaches that were validated for either condition alone.^51^ The VA and DoD have issued guidelines for managing insomnia and OSA, with 41 recommendations,^52,53^ yet no guidance currently exists for treating COMISA specifically, and guidelines are also not available from the American Academy of Sleep Medicine. Notably, combined treatment using cognitive behavioral therapy for insomnia (CBTi) alongside CPAP has shown greater efficacy than sequential or single-modality interventions.^46–48,54^ While CBTi is a promising technique for COMISA management, caution is warranted with non-selective use of CBTi components. For instance, a retrospective study of Veterans found reduced benefit from brief behavioral therapy among non-White patients and those with a short sleep duration (<4.1 hours).^55^ When medications are appropriate, newer non-benzodiazepine sedative hypnotics and anti-depressant agents could be helpful among patients with COMISA.^47^ Adjunctive strategies such as sleep hygiene and education are also supported.^56^ Importantly, treatment should be tailored to sex-specific symptom profiles and preferences. Women are less likely to report classic OSA symptoms,^57^ and are more likely to prefer non-pharmacological treatments.^58^ Additionally, cross-sectional survey data from 1,141 post-9/11 Veterans (53% women) showed that while women were more likely to endorse sleep as a behavior that they believed could prevent CVD,^59^ men and women Veterans are equally unlikely to endorse “getting adequate sleep” as part of their CVD prevention. Acknowledging such differences and similarities can enhance detection and education, and optimize individualized care.^60^

There are notable strengths to this investigation. Our cohort allowed for robust estimation of risks and subgroup analyses, especially differences between men and women. The 20-year follow-up also offers a strong temporal relation between sleep disorders and incident cardiovascular outcomes, making causality more plausible. Also, highlighting the elevated cardiovascular risk among younger adults and emphasizing early detection and treatment of sleep disorders is clinically meaningful and potentially practice changing. Using administrative and electronic health record data and rigorous statistical methods including sensitivity analyses strengthens the validity of the findings.

There are also limitations. First, the sample was limited to post-9/11 Veterans receiving VA care, which may reduce generalizability to other Veteran cohorts, non-VA users, or the general population due to unique psychosocial characteristics. Replication in non-VA populations is warranted. Second, hypertension may be underdiagnosed among younger Veterans and women.^61,62^ To ensure that the hypertension estimates were conservative, patients were not included in the primary outcome based on qualifying BP measurements alone, although the results were similar when outpatient BP readings were also used to define hypertension. Third, as this retrospective cohort study involved EHR data, there are risks of selection bias, misclassification, and missing data. Since insomnia is underdiagnosed in the VA,^10^ results may be disproportionately attributable to OSA, and it is necessary to reevaluate the effects of COMISA in Veterans who participated in a comprehensive sleep evaluation, including objective measures and diagnostic severity for both OSA and insomnia, as well as treatment adherence. Relatedly, there was no information on the severity of OSA or insomnia, or CPAP adherence. Prospective assessments or validation studies could be suggested as future directions to confirm findings. Fourth, we could not assess insomnia phenotypes such as short sleep duration, which may be more strongly associated with hypertension and CVD risk,^63,64^ possibly leading to effect size underestimation. Fifth, residual confounding from unmeasured lifestyle factors – including sleep duration and quality, physical activity, diet, and stress – is likely and may vary by sex, race, ethnicity, and socioeconomic status.^65–67^ Sixth, indications for antihypertensive medications were not available. Finally, care received outside the VA was not captured.

In summary, both insomnia and OSA were independently associated with a significantly greater risk of incident hypertension and CVD among a sample of 1 million younger and middle-aged adults. COMISA – the co-occurrence of both conditions – was associated with the greatest risk: a over a 2-fold increase in hypertension risk, and elevated CVD risk of over 3-fold, respectively. These findings meaningfully strengthen the rationale for improved recognition of sleep disorders and support a stronger clinical focus on COMISA. While further research is needed to evaluate treatment effects and guide implementation of VA sleep care guidelines, early diagnosis and integrated management may significantly improve cardiovascular outcomes and quality of life for both men and women.

## Data Availability

The data are proprietary to the U.S. government.

## Non-standard Abbreviations and Acronyms

aHR: adjusted hazard ratio
BP: blood pressure
CI: confidence interval
CVD: cardiovascular disease
EHR: electronic health record
GAD: generalized anxiety disorder
HR: hazard ratio
ICD-9-CM and 10: International Classification of Diseases, Ninth Revision, Clinical Modification and Tenth Revision
MDD: major depressive disorder
OSA: obstructive sleep apnea
PTSD: posttraumatic stress disorder
VA: Veterans Health Administration

## Acknowledgements

We would like to thank all the Veterans for their service and contributions to this investigation. The views and opinions of authors expressed herein do not necessarily state or reflect those of the United States Government.

